# Prediction of Mortality in hospitalized COVID-19 patients in a statewide health network

**DOI:** 10.1101/2021.02.17.21251758

**Authors:** Bharath Ambale-Venkatesh, Thiago Quinaglia, Mahsima Shabani, Jaclyn Sesso, Karan Kapoor, Matthew B Matheson, Colin O Wu, Christopher Cox, Joao A C Lima

**Author notes:** Corresponding Author: Joao A. C. Lima, MD, Halsted 574, Cardiology Division, Department of Medicine, Johns Hopkins Hospital, 600 North Wolfe Street, Baltimore, MD, 21287.

## Abstract

**Importance:** A predictive model to automatically identify the earliest determinants of both hospital discharge and mortality in hospitalized COVID-19 patients could be of great assistance to caregivers if the predictive information is generated and made available in the immediate hours following admission.

**Objective:** To identify the most important predictors of hospital discharge and mortality from measurements at admission for hospitalized COVID-19 patients.

**Design:** Observational cohort study.

**Setting:** Electronic records from hospitalized patients.

**Participants:** Patients admitted between March 3^rd^ and August 24^th^ with COVID-19 in Johns Hopkins Health System hospitals.

**Exposures:** 216 phenotypic variables collected within 48 hours of admission.

**Main Outcomes:** We used age-stratified (<60 and >=60 years) random survival forests with competing risks to identify the most important predictors of death and discharge. Fine-Gray competing risk regression (FGR) models were then constructed based on the most important RSF-derived covariates.

**Results:** Of 2212 patients, 1913 were discharged (age 57±19, time-to-discharge 9±11 days) while 279 died (age 75±14, time to death 14±15 days). Patients >= 60 years were nearly 10 times as likely to die within 60 days of admission as those <60. As the pandemic evolved, the rate of hospital discharge increased in both older and younger patients. Incident death and hospital discharge were accurately predicted by measures of respiratory distress, inflammation, infection, renal function, red cell turn over and cardiac stress. FGR models for each of hospital discharge and mortality as outcomes based on these variables performed well in the older (AUC 0.80-0.85 at 60-days) and younger populations (AUC >0.90 at 60-days).

**Conclusions and Relevance:** We identified markers collected within 2 days of admission that predict hospital discharge and mortality in COVID-19 patients and provide prediction models that may be used to guide patient care. Our proposed model suggests that hospital discharge and mortality can be forecasted with high accuracy based on 8-10 variables at this stage of the COVID-19 pandemic. Our findings also point to several specific pathways that could be the focus of future investigations directed at reducing mortality and expediting hospital discharge among COVID-19 patients. Probability of hospital discharge increased over the course of the pandemic.

**Key Points:** *Question:* Can we predict the likelihood of hospital discharge as well as mortality from data obtained in the first 48 hours from admission in hospitalized COVID-19 patients?

*Findings:* Models based on extensive phenotyping mined directly from electronic medical records followed by variable selection, accounted for the competing events of hospital death versus discharge, predicted both death and discharge with area under the receiver operating characteristic curves of >0.80.

*Meaning:* Hospital discharge and mortality can be forecasted with high accuracy based on just 8-10 variables, and the probability of hospital discharge increased over the course of the pandemic.

## Introduction

The clinical spectrum of coronavirus disease 2019 (COVID-19) ranges from asymptomatic carriers who can transmit the virus to a mild clinical upper respiratory infection that can progress to acute respiratory distress syndrome with a high fatality rate.^1,2^ In addition to the upper respiratory disease presentation, direct inflammatory damage and microvascular thrombosis induced by COVID-19 can affect the tissue and vasculature of multiple organs in addition to the lungs, leading to severe life-threatening disease in a large number of affected individuals.^3,4^ Among those hospitalized, approximately 10-15% become critically ill requiring ICU monitoring and/or mechanical ventilation, and many succumb to multi-organ failure.^2,5^ Therefore, it is crucial that physicians be able to identify the predictors of vital organ failure and death.

However, from the perspective of patients being hospitalized for COVID-19 at the present time, the most important consideration is their likelihood of eventual discharge from the hospital. From this perspective, identifying predictions of hospital discharge to guide optimization of medical care is among the most crucial pieces of needed information. While they reflect the physicians’ imperative to efficiently recognize and stratify patient risk, predictors of hospital discharge and incident mortality may provide important complementary information, particularly if the predictive information is generated and made available in the immediate hours following admission, during the time window that largely defines COVID-19 disease outcomes.

We examined such a clinical predictive model constructed from data collected from all COVID-19 patients within 48 hours of admission to the five hospitals of one large health care system in the state of Maryland. We utilized a combination of machine learning guided variable selection coupled with established analytic methods to select the most important clinical markers and pathways that predict both time to discharge and incident mortality.

## Methods

### Patient population

The entire dataset was extracted from the Johns Hopkins Health System (JHHS) electronic medical record system, EPIC. JHHS has a diverse referral base, including the city of Baltimore as well as surrounding suburban areas adjacent to Washington DC in Maryland and neighboring states. The institutional review boards of all five affiliated hospitals approved this study and waived requirements for individual informed consent. All patients consecutively admitted with confirmed COVID-19 between March 3 and August 24, 2020 were included. SARS-CoV-2 was detected by using RT-PCR assays with the vast majority of samples were collected via nasopharyngeal swabs. Time to hospital discharge or death were considered as competing primary outcomes for the study.

Measurements collected included demographics, social history, comorbidities, and ancillary studies. Clinical parameters encompassed vital signs, encounter diagnosis and clinical problems listed at presentation. All measurements entered in the analysis were collected within 48 hours of admission and only those available in at least 50% of the patient population were included. Overall, 216 features obtained from diverse domains were utilized in the analysis. Figure S1 (supplementary Figure 1) shows the number of missing variables per patient of the variables that were included in the analysis. The supplement includes more details on the measurements.

### Statistical Analysis

The distributions of variables were evaluated to determine population data structure and assess skewness. Logarithmic transformations were used for variables with skewed distributions. For time-to-event analyses, the origin (t_0_) was the time of admission. Missing values were assumed to be missing at random, and random forest imputation was used as previously described.^6^

We used random survival forests (RSF) with competing risks,^7^ including death and hospital discharge as competing events for efficient variable selection and identification of the most important parameters associated with time to the two events of interest. The splitting rule was based on Gray’s test for improved prediction of cumulative event incidence.^8^ The most important event-specific predictors were identified using variable importance from permutation (VIMP).^9^ A backward stepwise competing risk Fine-Gray regression model with minimization of the Bayesian Information Criterion starting from the top-20 RSF predictors was generated for each endpoint to estimate the cumulative incidence function (CIF) for each event of interest. The time-dependent area under the receiver-operator curves based on each parsimonious Fine-Gray model were then calculated.^7,10–13^ Further details on model development, validation and statistical methods are provided in the Supplement.

## Results

Between March 3^rd^ and August 24^th^ 2020, 2212 patients were hospitalized with a diagnosis of COVID-19 at one of the five JHHS-affiliated hospitals. Of these, 1913 were discharged while 279 died while in the hospital. The mean (SD) time to discharge was 9±11 days while the mean time to death during hospitalization was 14±15 days.

Age showed significant non-linearity (Figure S3), where in participants over 60 years, greater age was linearly associated with greater chance of death and lower chance of discharge. In those below 60 years, age was associated with both outcomes with much lower slopes. Given this observation, the significant interactions between age and most covariates as well as comorbidities, the analyses were performed separately in participants below and at-or-above 60 years of age. Table 1 provides patient characteristics for the entire population stratified by those older and younger than 60 years of age, and Figure 1 shows the overall CIF for hospital death and discharge over time as calculated from the RSF analysis.

**Figure 1:**
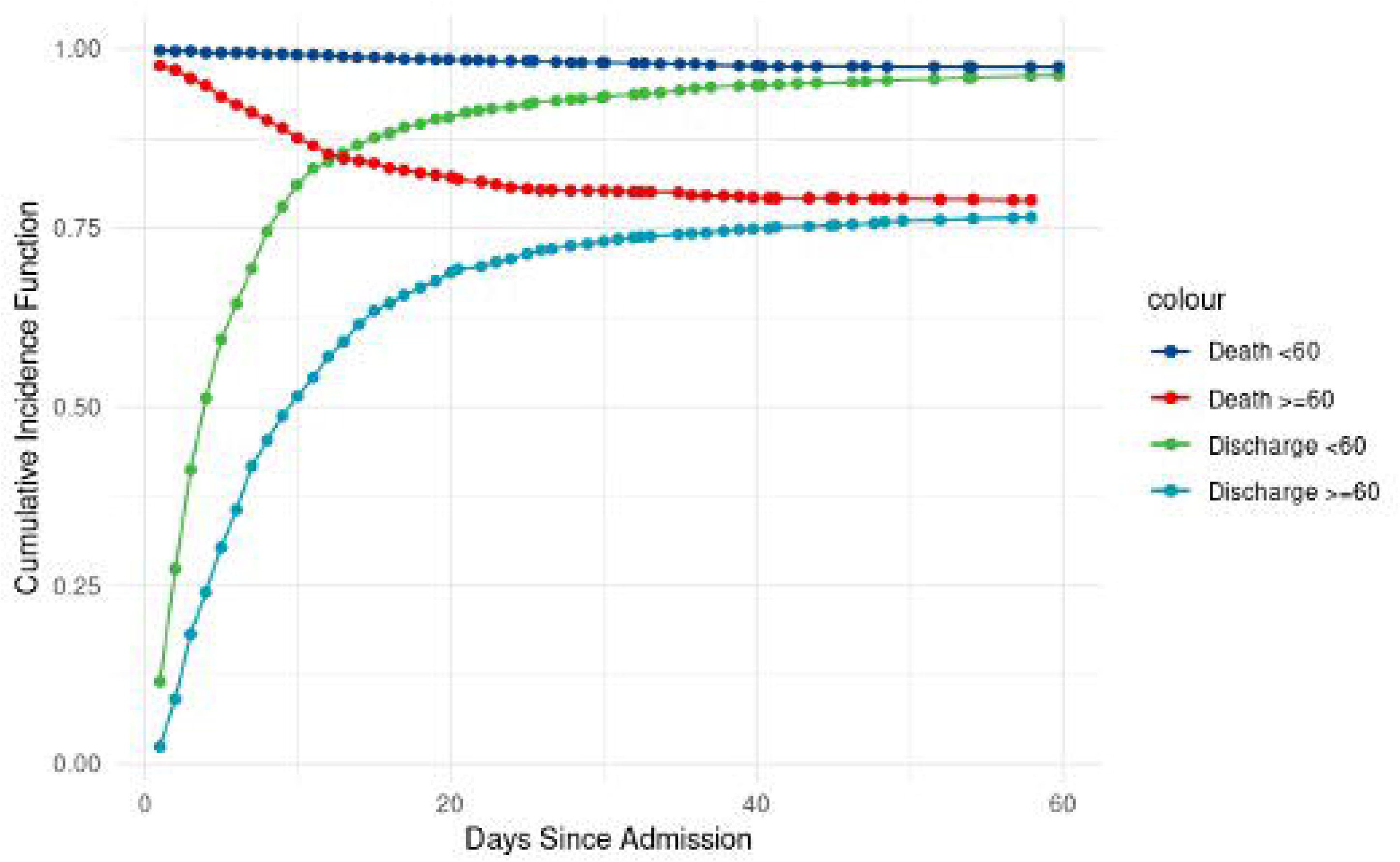
Cumulative Incidence Function (CIF) for death and discharge from the competing risk Random Survival Forests models for the patients under 60 years of age and patients 60 and over. The curves for death are represent 1-CIF. * - The number of deaths in the younger age group was n=30, appropriate care should be taken when interpreting the results.

**Table 1:** Patient characteristics at admission (only those assessed within 48 hours of admission) by outcome (death and discharge) in the old (60 years and over) and young (<60 years). Values are provided as mean ± standard deviation when continuous and normally distributed, median (inter-quartile range) when skewed, and as % when categorical.

| | Age ≥60 years | | Age <60 years | |
| --- | --- | --- | --- | --- |
| Characteristics | Discharged<br>(n=852) | Death<br>(n=249) | Discharged<br>(n=1061) | Death<br>(n=30) |
| Time to Event | 11±13 | 13±15 | 7±10 | 21±17 |
| Age in Years | 74±10 | 78±10 | 43±11 | 49±8 |
| Gender, female (%) | 49.9 | 45.8 | 46.4 | 20 |
| Hispanic ethnicity, % | 13.6 | 9.7 | 44.6 | 27.6 |
| Race, % |  |  |  |  |
| Other | 12 | 8.6 | 41.4 | 27.6 |
| Asian | 6.7 | 8.2 | 4.9 | 0 |
| Black | 41.6 | 35.2 | 34.3 | 58.6 |
| Indian | 0.1 | 0 | 0.5 | 0 |
| Pacific Islander | 0 | 0 | 0.3 | 0 |
| White | 39.6 | 48 | 18.7 | 13.8 |
| Body mass index, % |  |  |  |  |
| Normal | 37.8 | 47.3 | 16.2 | 16.7 |
| Overweight | 31.2 | 27.9 | 30.1 | 33.3 |
| Class 1 obesity | 16.4 | 14.9 | 26.8 | 20.8 |
| Class 2 obesity | 9 | 5.5 | 13.1 | 4.2 |
| Class 3 obesity | 5.7 | 4.5 | 13.8 | 25 |
| Oxygen support device used, % |  |  |  |  |
| High Flow Nasal Cannula | 56.7 | 20.9 | 68 | 16.7 |
| Mask | 2.2 | 7.8 | 2.4 | 23.3 |
| Nasal Cannula | 3.7 | 16.5 | 1.8 | 6.7 |
| Non-invasive positive pressure ventilation | 35.4 | 40.4 | 25.7 | 36.7 |
| Room air | 0.5 | 2.6 | 0.3 | 3.3 |
| Other, unspecified | 0.4 | 5.2 | 0.3 | 0 |
| Tracheal Collar | 0.1 | 0.4 | 0 | 0 |
| Mechanical Ventilator | 1 | 6.1 | 1.6 | 13.3 |
| Days Since Start of Pandemic | 65±32 | 54±24 | 69±33 | 59±25 |
| Pulse, beats per minute | 84±17 | 94±23 | 89±16 | 103±22 |
| Respiratory rate, breaths per minute | 21±6 | 25±8 | 21±5 | 25±6 |
| Body Temperature in C | 37.2±0.7 | 37.2±1.1 | 37.3±0.8 | 37.7±0.9 |
| Pulse oxygen saturation, % | 96.3±2.8 | 94.7±6.1 | 96.6±3.0 | 96.0±2.6 |
| Systolic blood pressure, mmHg | 129±22 | 120±26 | 123±20 | 120±24 |
| Hemoglobin, g/dL | 12.7 (2.6) | 12.4 (3.5) | 13.4 (2.6) | 13.0 (3.2) |
| Mean Corpuscular Volume, fL | 89.3 (8.0) | 91.6 (2.0) | 86.9 (6.6) | 88.2 (10.2) |
| Mean corpuscular hemoglobin concentration, g/dL | 32.5 (2.0) | 32.0 (2.0) | 33.0 (1.8) | 31.8 (2.5) |
| Red Cell Distribution Width, % | 13.8 (1.9) | 14.8 (3.0) | 13.2 (2) | 14.6 (1.9) |
| Pro B-type Natriuretic Peptide, mg/dL | 401 (1550) | 1176 (3835) | 51 (146) | 391 (1350) |
| Blood urea nitrogen, mg/ dL | 20.0 (18.0) | 32.0 (33.0) | 10 (7) | 22(19) |
| Creatinine, mg/dL | 1.11 (0.74) | 1.41 (1.20) | 0.84 (0.39) | 1.41 (2.24) |
| Aspartate Aminotransferase, /L | 38.0 (30.0) | 43.0 (43.0) | 38 (35) | 62 (43) |
| Alanine Aminotransferase, /L | 25.0 (25.0) | 27.0 (27) | 36 (40) | 36 (44) |
| AST/ALT ratio | 1.51 (0.80) | 1.71 (0.90) | 1.11 (0.50) | 1.61 (1.35) |
| C-reactive protein, mg/dL | 11.5 (29.2) | 44.6 (114) | 6.9 (14.6) | 17.8 (18.3) |
| D-Dimer, mcg/mL FEU | 1.1 (1.5) | 2.2 (3.5) | 0.7 (0.8) | 2.2 (2.2) |
| Procalcitonin, ng/mL | 0.2 (0.5) | 0.5 (1.9) | 0.15 (0.23) | 0.70 (1.10) |
| White blood cell count, /nL | 6.8 (4.3) | 8.1 (6.1) | 6.7 (4.0) | 8.3 (5.5) |
| Eosinophil, % | 0.1 (0.6) | 0 (0.2) | 0.1 (0.6) | 0 (0.3) |
| Neutrophil, % | 75.2 (15.5) | 80.9 (11.9) | 73.3 (15.0) | 79.8 (8) |
| Monocyte, % | 7.7 (4.7) | 6.3 (4.9) | 6.6 (4.1) | 5.5 (3.5) |
| Ferritin, mg/dL | 604 (807) | 894 (1342) | 510 (916) | 856 (802) |
| Lactate Dehydrogenase, /L | 334 (233) | 477 (322) | 323 (206) | 484 (366) |
| Creatine Kinase, /L | 128 (204) | 214 (312) | 114 (180) | 511 (447) |
| **Comorbid Conditions, %** |  |  |  |  |
| Chronic lung disease | 20 | 23.8 | 17 | 20.7 |
| Blood loss anemia | 1.2 | 1.7 | 3 | 0 |
| Psychoses | 7.2 | 10.4 | 5 | 10.3 |
| Peripheral vascular disease | 12.4 | 17.1 | 2.2 | 3.4 |
| Pulmonary circulation disorder | 5.8 | 6.7 | 3.9 | 3.4 |
| Hypothyroidism | 11.5 | 14.2 | 4.5 | 6.9 |
| Neurological disorders | 27 | 42.5 | 9.1 | 24.1 |
| Deficiency anemias | 23.7 | 32.1 | 16.1 | 20.7 |
| Renal failure | 18.8 | 26.7 | 6.5 | 34.5 |
| Liver disease | 5.4 | 3.3 | 6.1 | 6.9 |
| Arthritis | 4.4 | 4.6 | 2.8 | 3.4 |
| Diabetes with complications | 19.6 | 18.3 | 12 | 27.6 |
| Solid tumor without metastasis | 14 | 19.2 | 2.5 | 3.4 |
| Depression | 21.3 | 27.1 | 12.7 | 20.7 |
| AIDS | 1.2 | 0.4 | 1.7 | 3.4 |
| Hypertension with complications | 7.2 | 7.1 | 3.3 | 10.3 |
| Coagulation deficiency | 4.9 | 7.1 | 3.8 | 0 |
| Metastatic cancer | 7.3 | 10.4 | 2.6 | 3.4 |
| Heart failure | 19.6 | 25.4 | 7.5 | 27.6 |
| Paralysis | 3.3 | 5.8 | 0.9 | 0 |
| Fluid and electrolyte disorders | 33.8 | 41.7 | 14.8 | 20.7 |
| **Problems list at Admission, %** |  |  |  |  |
| Abnormal Chest Xray | 0.4 | 0.4 | 0.6 | 3.3 |
| Abnormal ECG | 0.6 | 0.8 | 0.5 | 0 |
| Acute coronary syndrome | 2.3 | 5.2 | 1.1 | 3.3 |
| Acute HF | 4.6 | 5.6 | 1.7 | 3.3 |
| Acute kidney injury | 9.9 | 17.7 | 3 | 16.7 |
| Acute pulmonary embolism | 2.2 | 2.8 | 1.8 | 0 |
| Anosmia | 0.8 | 0.8 | 0.2 | 0 |
| Cardiac arrest | 1.3 | 2.4 | 0.3 | 3.3 |
| Chest pain | 4.2 | 1.2 | 5.8 | 0 |
| Cough | 3.8 | 1.2 | 4 | 0 |
| Delirium | 1.4 | 0.8 | 0.3 | 0 |
| Diarrhea | 2.1 | 2.4 | 1.2 | 3.3 |
| Nausea | 1.3 | 2.4 | 2.9 | 0 |
| Exacerbation of COPD or Asthma | 0.6 | 1.2 | 1.9 | 0 |
| Fever or Chills | 7.3 | 8.4 | 7.3 | 3.3 |
| Headache | 0.6 | 0.4 | 1.5 | 0 |
| Hypoxia, % | 15.8 | 16.9 | 13.2 | 23.3 |
| Fatigue | 7.4 | 4 | 1.5 | 0 |
| Palpitation/Arrhythmia | 3.8 | 1.2 | 4.6 | 6.7 |
| Pneumonia | 31.5 | 36.5 | 26.9 | 23.3 |
| Sepsis, % | 5.8 | 18.5 | 1.9 | 13.3 |
| Shock | 0.8 | 4.4 | 0.6 | 0 |
| Stroke | 2.5 | 2.8 | 0.8 | 0 |

### Evolution of prognosis for patients hospitalized over the time course of the pandemic

To account for the evolving nature of medical therapy and prognosis over the time course of the pandemic, we included the time from the start of the pandemic in the state of Maryland (time of first admission to the JHHS system - March 3^rd^), relative to each patient’s date of admission as a covariate. Figure 2 shows the partial effect of time since the local pandemic start to the CIF calculated at 60 days, taking into account the effect of all covariates in the RSF predictive model. The figure shows the progressive decrease of the CIF for death, with corresponding CIF increase for hospital discharge at 60 days after admission, as the pandemic evolved. The magnitude of changes in the CIFs for mortality were of the order of 20-25% in both the older and younger patients. Consequently, all models included this as a covariate.

**Figure 2.**
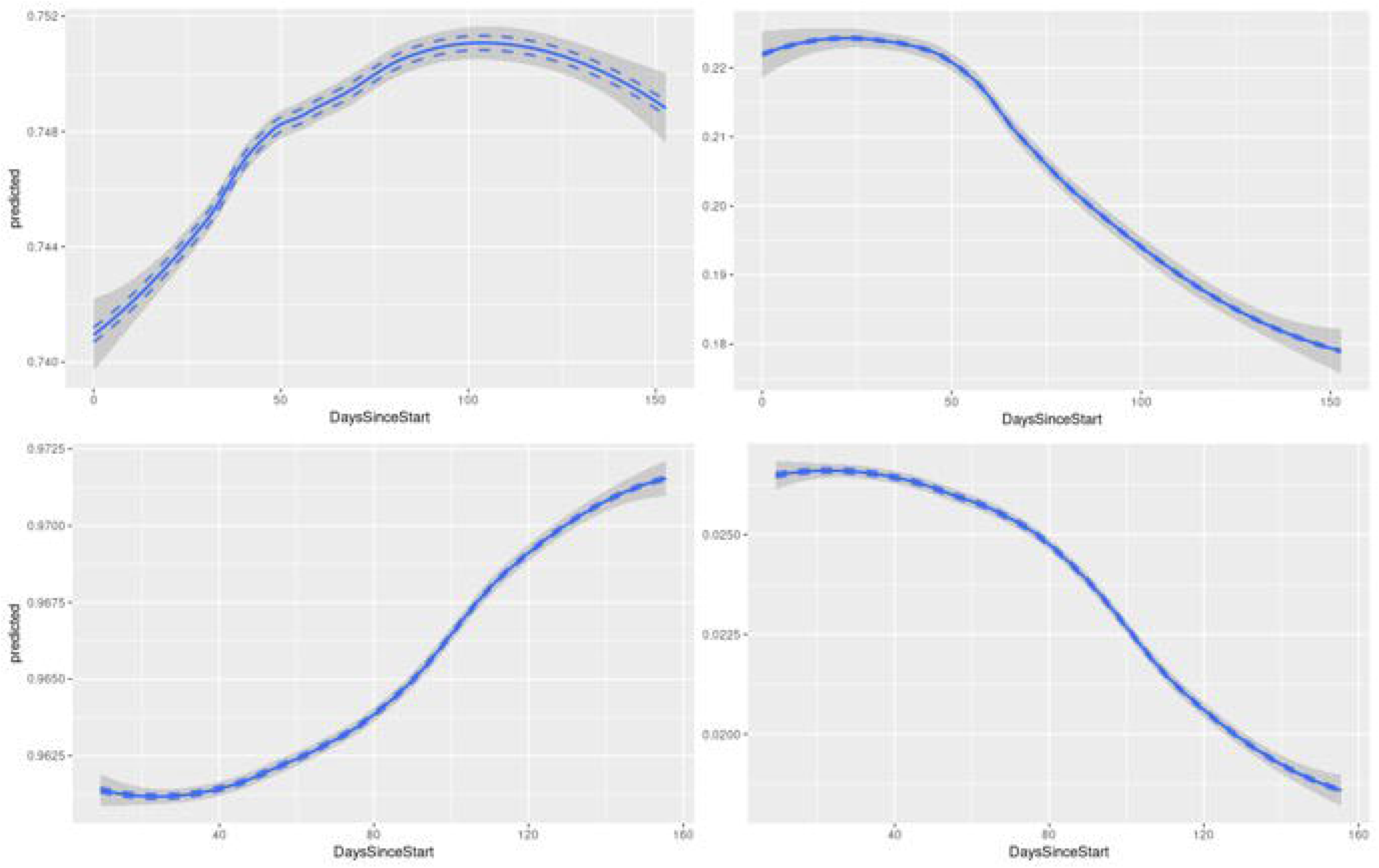
Partial variable dependence plots (with loess smoothing) showing the association of time since the start (x-axis) of the pandemic in Maryland (since March 3^rd^) with the cumulative incidence function at 60 days post-admission (y-axis) for discharge (left column) and death (right column) as computed from the competing risk Random Survival Forests model with all variables included in the >=60 year old (top row) and <60 year old patients (bottom row). * - The number of deaths in the younger age group was n=30, appropriate care should be taken when interpreting the results.

### Variable importance calculated from RSF

The most important predictors of hospital discharge and death are shown in Table S1 in descending order of importance. Thirty among the 279 patients who died while hospitalized were younger than 60 years. As a consequence, predictive power for death among patients younger than 60 years was limited, and the corresponding results should be interpreted with caution.

Figure S4 displays the relative importance for all variables across different domains pertaining to hospital discharge and mortality for patients 60 years and older (top) as well as to hospital discharge among those younger than 60 years of age (bottom). When compared to breathing room air, the magnitude of oxygen requirement in the first 48 hours from admission expressed as flow rate, device complexity and invasiveness, progressively reduced the likelihood and increased the time to hospital discharge among older and younger patients, and predicted death among those aged 60 and older. Lower respiratory and heart rates, lower BUN and procalcitonin levels, lesser inflammation as indexed by lower CRP levels, lower eosinophil count as well as reduced red cell distribution width (RDW) were associated with greater likelihood and shortened time to hospital discharge in both the 60 and older and younger than 60 years of age groups. The same were associated with opposite effects for incident death among those 60 and older. Other measures featured prominently on the specific lists of most important predictors for the two outcomes in both age groups are shown in Table S1 and Figure S4. Supplemental figures S5-S8 show partial effect plots from the RSF model used to examine non-linear associations and directionality.

### Final prediction models for hospital discharge and mortality

Fine-Gray models were validated (Figure S9) and the performance of the parsimonious Fine-Gray models was similar to that of the RSF models with all variables (Figure S10) for each outcome of interest. The corresponding final model sub-distribution hazard ratios and p-values are shown in Table 2 for the older and younger age groups respectively.

**Table 2:** Coefficients (and associated p-values) from Fine-Gray competing risk regression models for prediction of (a) discharge (n=852) and death (n=249) after variable selection using RSF in patients 60 years of age and over; and (b) discharge (n=1061) and death (n=30) after variable selection using RSF in patients under 60 years of age. * - The number of deaths in the young population was n=30, therefore appropriate care should be taken when interpreting the results.

| **PATIENTS >=60 YEARS** |  |  |
| --- | --- | --- |
| **Endpoint: Discharge** | **Hazard Ratio** | **P-Value** |
| Log (C-reactive protein) | 0.86 | <0.001 |
| Respiratory rate | 0.98 | 0.007 |
| Log (Blood Urea Nitrogen) | 0.83 | 0.002 |
| Log (Red Cell Distribution Width) | 0.30 | <0.001 |
| Age (per decade) | 0.86 | <0.001 |
| Time since start of pandemic (per month) | 1.19 | <0.001 |
| Log (Absolute Eosinophil count) | 1.20 | <0.001 |
| Pulse rate | 0.99 | <0.001 |
| Sepsis | 0.53 | <0.001 |
| Oxygen device used |  |  |
| High flow nasal cannula | 0.28 | <0.001 |
| Mask | 0.37 | <0.001 |
| Nasal Cannula | 0.64 | <0.001 |
| Non-invasive positive pressure ventilation | 0.29 | 0.003 |
| Other | 0.15 | <0.001 |
| Tracheal Collar | 1.27 | 0.85 |
| Mechanical Ventilator | 0.27 | <0.001 |
| **Endpoint: Death** | **Hazard Ratio** | **P-Value** |
| Respiratory rate | 1.04 | <0.001 |
| Log (Red Cell Distribution Width) | 10.52 | <0.001 |
| Age (per decade) | 1.35 | <0.001 |
| Time since pandemic start (per month) | 0.75 | <0.001 |
| Log (Mean Corpuscular Volume) | 9.93 | 0.009 |
| Log (C-reactive protein) | 1.37 | <0.001 |
| Pulse rate | 1.02 | <0.001 |
| Systolic Blood Pressure (per 10 mm Hg) | 0.90 | 0.003 |
| **PATIENTS <60 YEARS** |  |  |
| **Endpoint: Discharge** | **Hazard Ratio** | **P-Value** |
| Age (per decade) | 0.84 | <0.001 |
| Time since pandemic start (per month) | 1.13 | <0.001 |
| Respiratory rate | 0.97 | <0.001 |
| Log (Creatinine) | 0.80 | 0.002 |
| Log (Procalcitonin) | 0.84 | <0.001 |
| Log (pro B-type natriuretic peptide) | 0.95 | 0.085 |
| Log (Red Cell Distribution Width) | 0.36 | <0.001 |
| Pulse rate | 0.99 | <0.001 |
| Log (Ferritin) | 0.93 | 0.021 |
| Oxygen device used |  |  |
| High flow nasal cannula | 0.39 | <0.001 |
| Mask | 0.35 | <0.001 |
| Nasal Cannula | 0.70 | <0.001 |
| Non-invasive positive pressure ventilation | 0.31 | 0.004 |
| Other | 0.50 | 0.26 |
| Mechanical Ventilator | 0.32 | <0.001 |
| **Endpoint: Death\*** | **Hazard Ratio** | **P-Value** |
| Pulse rate | 1.03 | 0.003 |
| Log (Blood Urea Nitrogen) | 1.63 | <0.001 |
| Log (Procalcitonin) | 2.81 | <0.001 |
| Log (AST/ALT ratio) | 5.80 | <0.001 |

Among hospitalized patients older than 60 years of age, hospital discharge could be predicted with an area under the curve of 0.84 (CI:0.81 - 0.87) at 60 days (Figure 3) by a combination of 10 variables: 1) lower respiratory rate, 2) lower CRP, 3) younger age, 4) lower heart rate, 5) longer time since the start of the pandemic, 6) absence of sepsis, 7) lower BUN, 8) less altered RDW, 9) higher eosinophil count and 10) type of oxygen device used in the first 48 hours following hospital admission. Similar variables predicted mortality among hospitalized patients aged 60 years of age and older with an AUC of 0.80 (CI: 0.77-0.83). They included: 1) higher respiratory rate, 2) greater RDW, 3) greater age, 4) shorter time since the pandemic started, 5) higher heart rate, 6) greater mean corpuscular volume, 7) higher C-reactive protein, and 8) lower systolic blood pressure in the first 48 hours from admission.

**Figure 3:**
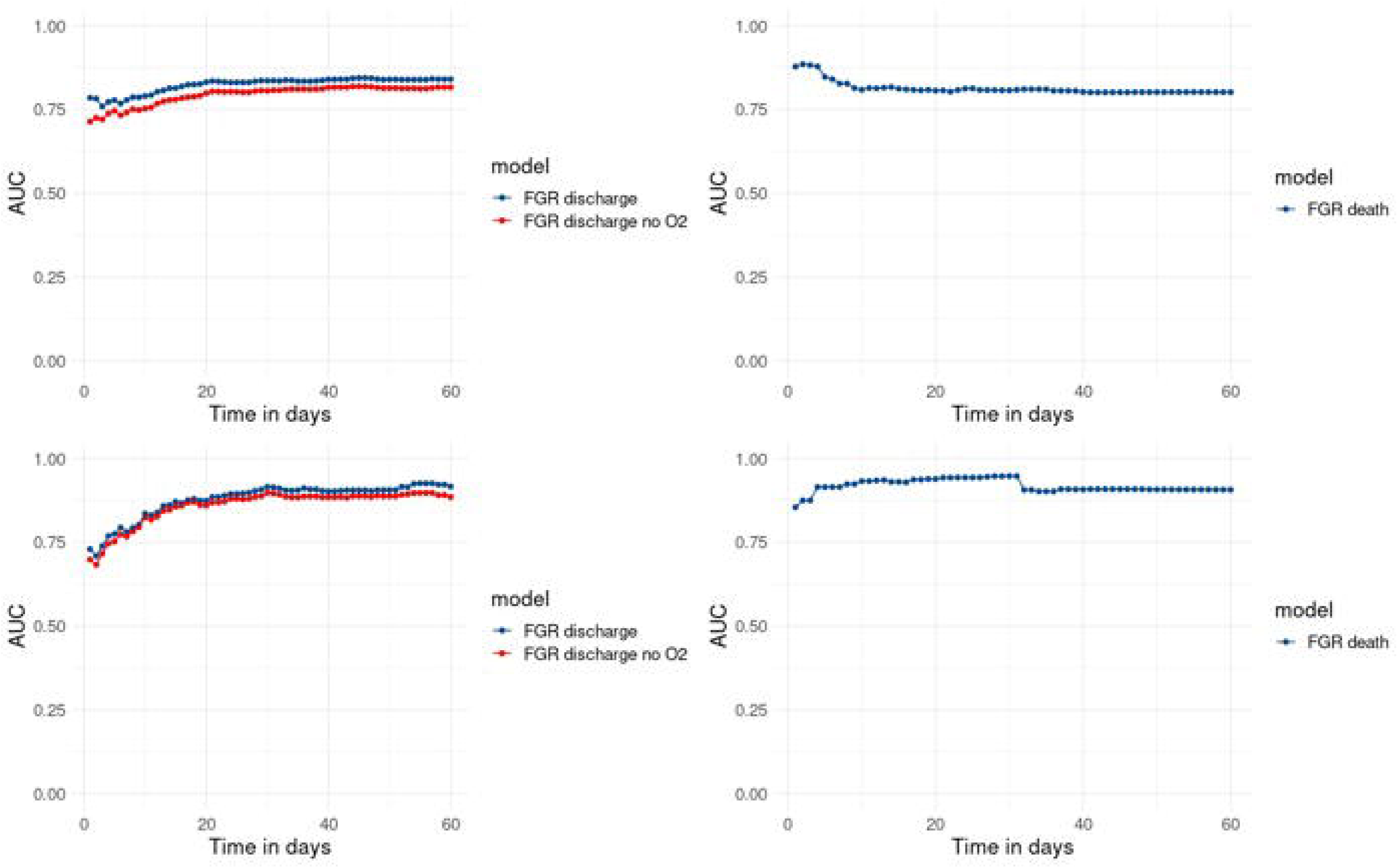
Time-dependent area under the ROC for discharge (left panels) and death (right panels) from the competing risk Fine-Gray regression (FGR) models in patients 60 years and over (top row) and patients under 60 (bottom row). * - The number of deaths in the younger age group was n=30, appropriate care should be taken when interpreting the results.

Similarly, hospital discharge among COVID-19 patients younger than 60 could be predicted with AUC of 0.92 (CI:0.87 - 0.96) at 60 days by a combination of 10 slightly different variables: 1) younger age, 2) greater time since the start of the pandemic, 3) lower respiratory rate, 4) lower creatinine, 5) lower procalcitonin; 6) lower NTproBNP levels, 7) reduced RDW, 8) lower heart rate; 9) lower ferritin and 10) type of oxygen device used within the first 48 hours of hospital admission. Notwithstanding the limitations mentioned above, the predictors for death selected by the final model for patients younger than 60 years of age included: 1) greater AST/ALT ratio, 2) higher BUN, 3) higher procalcitonin levels, and 4) greater heart rate.

Finally, because oxygen device prescribed in the first 48 hours from admission could be considered both as a predictor and a treatment modality, we constructed models for likelihood and time to discharge excluding type of oxygen device. Predictive power was reduced by 3-4% on average for each model as shown in Figure 3.

## Discussion

The results of this study suggest that statistical learning methods can be used for meaningful variable selection from routine, clinically acquired medical data. Based on such selection, parsimonious models were derived to risk stratify hospitalized COVID-19 patients and accurately predict the likelihood and time to hospital discharge and death. Respiratory dysfunction phenotypes combined with selected parameters reflecting systemic inflammation, organ/tissue damage, metabolic dysregulation, red cell turnover and coagulation abnormalities predicted hospital discharge and death with AUC = 0.84 (CI:0.81 - 0.87) and AUC = 0.80 (CI: 0.77 - 0.83) respectively for patients older than 60 years, as well as hospital discharge with AUC = 0.92 (CI: 0.87 - 0.96) for those younger than 60 years of age. Our findings also suggest that as the pandemic evolved, the adjusted death rates dropped, while the probability and rates of hospital discharge improved for both older and younger COVID-19 patients.

Pulmonary inflammatory and respiratory involvement captured by the respiratory rate and the type of device prescribed to deliver oxygen in the first 48 hours after admission were the two most important predictors of hospital discharge for both older and younger than 60 years COVID-19 patients. Greater respiratory rate also predicted mortality among older and younger COVID-19 patients.^14^ Greater eosinophil count predicted hospital discharge while lower eosinophil count predicted death during hospitalization among patients 60 and older. Immunologic response mediated in part by cytokine release and resulting inflammation has been previously implicated in both disease severity and ability to respond to the viremic insult.^15^ In this regard, CRP as a systemic inflammatory marker, was more useful among patients 60 and older as a predictor of both hospital discharge and mortality than in patients younger than age 60. Conversely, procalcitonin, a hormone reflecting a heightened immunologic state was a more prominent predictor among the younger patients with COVID-19.^16^ Both CRP and pro-calcitonin have been consistently found to predict disease severity and mortality in COVID-19 patients. Since COVID-19 emergence, signs and symptoms of sepsis associated with SARS-CoV-2 virus infection have been reported as hallmarks of disease progression in critically ill patients, with septic shock indicated as a major determinant of mortality in a large proportion of hospitalized patients.^1^ In our study, sepsis was strongly associated with later and less likely hospital discharge in the older age group.

The striking relationship between fatality rate and age has been previously documented^17,18^ and our analysis demonstrated a steep increase beginning at age 60, which defined our cutoff for age stratification. While immunologic response is known to be age dependent, COVID-19 age dependence appears to be also related to systemic vascular aging given its proclivity for endothelial damage. While pulmonary involvement determines the need for hospitalization and the intensity of care required, pre-existing and superimposed endothelial injury appears to underlie organ system disease severity, including the pulmonary system. In our study, heart rate increase, unlike respiratory rate increase, more likely reflects systemic demand as opposed to direct myocardial injury, although the latter cannot be entirely ruled out in most patients. Similarly, hypotension in the first two days of admission portended a greater risk of death in patients 60 or older, likely reflecting a combination of systemic and cardiovascular pathology. Accurate rates of cardiac and vascular involvement among inpatient and outpatient COVID-19 patients remain difficult to quantify at present.^19–22^

Renal function, as the ability to keep up with the increased byproduct load of upregulated catabolism from infection, inflammation and damage is further impaired by direct kidney endothelial injury in COVID-19. In our study, the predictive power of accumulated BUN was remarkable in the final model for hospital discharge in patients 60 and older, as well as for death among patients younger than 60 years of age. In addition, among the latter, lower creatinine was a major marker of hospital discharge, while hepatic dysfunction was a predictor of mortality.^23–25^

Importantly also, red blood cell turnover measured as RDW was a marker of both hospital discharge and mortality among patients 60 years and older, and also of hospital discharge among those younger than 60 years of age. Among older patients, red cell turnover as an index of systemic marrow stress was also reflected by the association of mean corpuscular volume (MCV) to outcomes.^26,27^ These findings support the concept of thrombotic microangiopathy as a potential pathogenetic process in COVID-19.^28^

The limitations of the predictive model for hospital mortality among patients younger than 60 have been emphasized. In that group, 1061 patients were discharged and only 30 died while in the hospital. Therefore, the model for mortality should be interpreted with caution. Another limitation of our study is that data are derived from a statewide health system and their generalizability is therefore affected by population demographic and other regional differences. Data on disease presentation and history of medical conditions from the electronic health record may be incomplete. Similarly, post-discharge outcomes were not included in this analysis and those occurring outside our health care system may have been missed.

A strength of this study is the use of competing risk to assess outcomes after hospitalization for COVID-19. As observed in other studies as well as in our study population, the time to discharge is relatively short and the likelihood of discharge is high after hospitalization for COVID-19. Given this scenario, considering patients who were discharged at a given time point as right-censored when in-hospital death is the primary outcome, implicitly assumes that they have similar risk of dying from COVID-19, compared to those who are still at risk (i.e. still in the hospital) at that time point. In such a situation, assessment of competing risks provides a more accurate assessment of risks. The study of time to discharge also adds to our understanding of prolonged hospital stay due to concomitant complications.

Finally, this study clearly shows the importance of cohort effect on the probability of hospital mortality and mirrored elevation in the likelihood and time to hospital discharge. Determinants of such differences cannot be quantified from this analysis. Hypothesized contributors include the implementation of treatments documented as effective in COVID-19.^29^ Moreover, oxygen delivery strategies such as proning and optimal timing for mechanical intubation changed as the pandemic evolved. Other modifications in management strategies may not have been as obvious and remain unaccounted.

In conclusion, we identified specific markers at admission that predict time to hospital discharge and mortality in patients admitted with COVID-19 and provide prediction models that may be used to guide patient care.

## Data Availability

The Authors are responsible for the accuracy and integrity of the entire set of results and are willing to provide, after IRB consent, the datasets and code used in the analysis at any time as requested.

## Author Contributions

The study was designed by the principal investigators Bharath Ambale-Venkatesh and Joao A C Lima in collaboration with the other authors.

Data Collection: Bharath Ambale-Venkatesh, Mahsima Shabani, Thiago Quinaglia, Jaclyn Sesso, Karan Kapoor

Analysis and interpretation of data: All authors.

Drafting of the manuscript: Bharath Ambale-Venkatesh and Joao A C Lima.

Critical revision of the manuscript for important intellectual content: All authors.

Statistical Analysis: Bharath Ambale-Venkatesh, Matthew Matheson, Colin Wu and Christopher Cox Administrative, technical or material support: Jaclyn Sesso

## Acknowledgements

We would like to thank the Johns Hopkins CROWN Program and the CADRE committee for making available the data used in this analysis and for curation and supervision of the data collection from COVID-19 patients admitted to the Johns Hopkins Healthcare System. We would also like to thank Jason Ortman, IT manager, for maintenance of the data storage and analytic environments.

