## Supplement for "Prediction of Mortality in hospitalized COVID-19 patients in a statewide health network"

### *Patient population*

The entire dataset was extracted from the Johns Hopkins Health System (JHHS) electronic medical record system, EPIC. JHHS has a diverse referral base, including the city of Baltimore as well as surrounding suburban areas adjacent to Washington DC in Maryland and neighboring states. The five JHHS-affiliated hospitals include: Johns Hopkins Hospital (Baltimore, MD), Johns Hopkins Bayview Medical Center (Baltimore, MD), Howard County General Hospital (Columbia, MD), Sibley Memorial Hospital (Washington, D.C), and Suburban Hospital (Bethesda, MD). We used the Precision Medicine Analytics Platform which pulls data from the EPIC Medical Record across the JHHS, where the data are integrated and made available in an operable format. This secure dataset and analytic environment is managed by the COVID-19 And Data Research Evaluation committee composed of JHHS providers, investigators and data scientists. The institutional review boards of all five affiliated hospitals approved this study and waived requirements for individual informed consent. All patients consecutively admitted with confirmed COVID-19 between March 3 and August 24, 2020 were included. SARS-CoV-2 was detected by using RT-PCR assays targeting different genes (e.g. N, E, or S proteins). The vast majority of samples were collected via nasopharyngeal swab although alternative confirmatory tests were also utilized including bronchoalveolar lavage, saliva and sputum. Time to hospital discharge or death were considered as competing primary outcomes for the study.

### *Measurements*

Measurements collected included demographics, social history, comorbidities, and ancillary studies. Clinical parameters encompassed vital signs, encounter diagnosis and clinical problems listed at presentation. All measurements entered in the analysis were collected within 48 hours of admission and only those available in at least 50% of the patient population were included. Overall, 216 features obtained from diverse domains were utilized in the analysis. Figure S1 shows the number of missing variables per patient of the variables that were included in the analysis.

Patient demographics, detailed medical history, comprehensive laboratory biomarkers, and summary findings from electrocardiograms and chest X-rays interpreted by attending cardiologists and radiologists respectively, were included in the model. JHHS uses text analytics algorithms to analyze medical records and reports, extracting individual key words and phrases. Using such techniques, features were extracted from high-dimensional text datasets. These features were corroborated by two clinicians and a practicing nurse (MS, TQ, JS), and a curated clinical problems list was created and coded into a format suitable for statistical analysis. The entire process incorporated data previously collected from patient exposure to COVID-19 both locally and elsewhere.

Laboratory data collected from each patient included complete blood count, liver enzyme and function tests, renal function tests and electrolytes (blood urea nitrogen - BUN, creatinine, glomerular filtration rate - GFR), cardiac disease markers (pro B-type natriuretic peptide - NTproBNP, troponin levels), lactate dehydrogenase - LDH, C-reactive protein - CRP, ferritin, and coagulation markers. Only biomarkers collected within the first 48 hours of admission were used in the analysis. Arterial blood gases included CO2 and anion gap. The FiO2 was defined according to O2 delivery (room air, nasal cannula, simple face mask, venturi mask, non-rebreather mask, high flow nasal cannula, or for mechanical ventilation according to input settings).

Figure S1: Graph illustrating the percent of missing datapoints across the study population. The y-axis denotes the number of patients. The x-axis shows the associated number of missing variables out of a total of 216 variables.

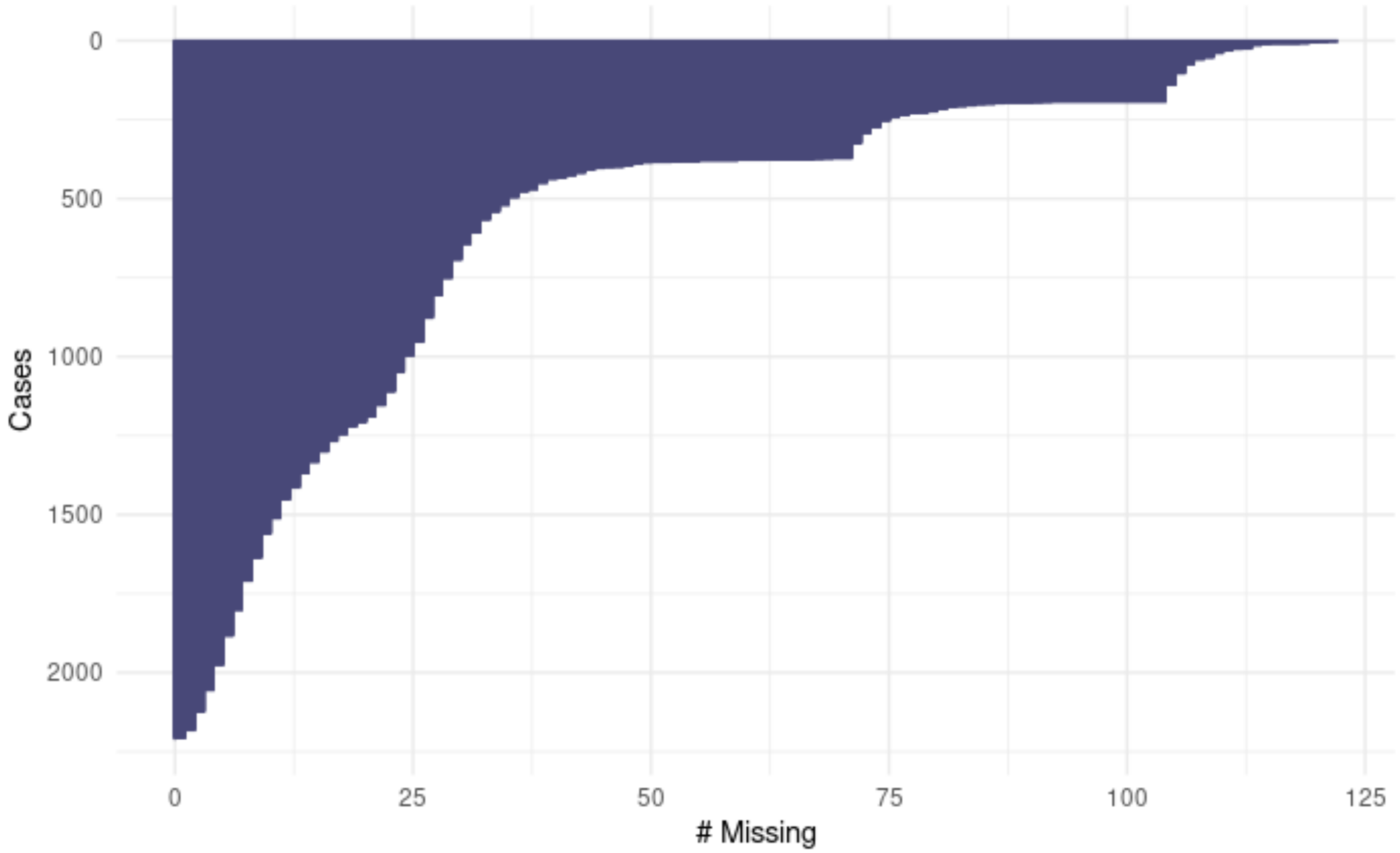

*STATISTICAL ANALYSIS*

The distributions of variables were evaluated to determine population data structure and assess skewness. Continuous variables were evaluated by Shapiro-Wilk or Kolmogorov-Smirnov tests for normality. Quantitative variables were reported as mean ± standard deviation OR median and interquartile range (IQR) if skewed, while qualitative variables were shown as frequency (%). Logarithmic transformations were used for variables with skewed distributions. For time-to-event analyses, the origin ($t_0$) was the time of admission. Missing values were assumed to be missing at random, and random forest imputation was used as previously described.[6]

We used random survival forests (RSF) with competing risks,[7] including death and hospital discharge as competing events for efficient variable selection and identification of the most important parameters associated with time to the two events of interest. Once the features from the data sources had been collected and transformed as detailed above, the RSF algorithm was applied for variable selection. Hyperparameter tuning was performed using a random search algorithm to identify the optimal RSF parameters. The splitting rule was based on Gray's test for improved prediction of cumulative event incidence.[8] Prediction error was calculated from out-of-bag (OOB) samples. The addition of trees was monitored to ensure OOB error stabilization and assure model convergence (Figure S2).

The most important event-specific predictors were identified using variable importance from permutation (VIMP).[9] Based on the top variables identified from RSF, a parsimonious Fine-Gray competing risk regression model[10] was constructed to estimate the cumulative incidence function (CIF) for each event of interest. The functional forms of the associations for each variable with the outcomes were estimated from partial effect plots obtained from RSF analysis.[7] We tested each covariate for proportionality; in the presence of proportional subhazards, the subhazard ratio can be interpreted as an effect on the relative risk of the event occurring. For example, when time to discharge is being modeled, a subhazard ratio above 1 indicates an increased hazard of discharge, thus making this event more likely for those with a higher value of the given predictor.

A backward stepwise competing risk Fine-Gray regression model with minimization of the Bayesian Information Criterion starting from the top-20 RSF predictors was generated for each endpoint. Model development was performed on a randomly chosen training subset (70% of the population) and applied on the test subset (remaining 30%) to ensure model calibration, validity and performance against the RSF models. The final model sub-distribution hazard ratios and corresponding confidence intervals and p-values are reported based on the entire population. The time-dependent area under the receiver-operator curves based on each parsimonious Fine-Gray model were then calculated.[11,12] Fine-Gray competing risk regression model validation showed that our models were well-calibrated with the predicted risk associated with the observed event frequencies in the test subset (Figure S9).

Data analysis was performed with R software using publicly available libraries for Fine-Gray regression, RSF methods, and time-varying receiver-operator curves (ROC) using the R packages randomForestSRC, ggRandomForest, crrstep, cmprsk, and riskRegression.[7,12–14]

Figure S2: Out-of-bag error with addition of trees in the building of the RSF for patients <60 (left) and >=60 years (right) showing both the models converged. * - The number of deaths in the younger age group was n=30, appropriate care should be taken when interpreting the results.

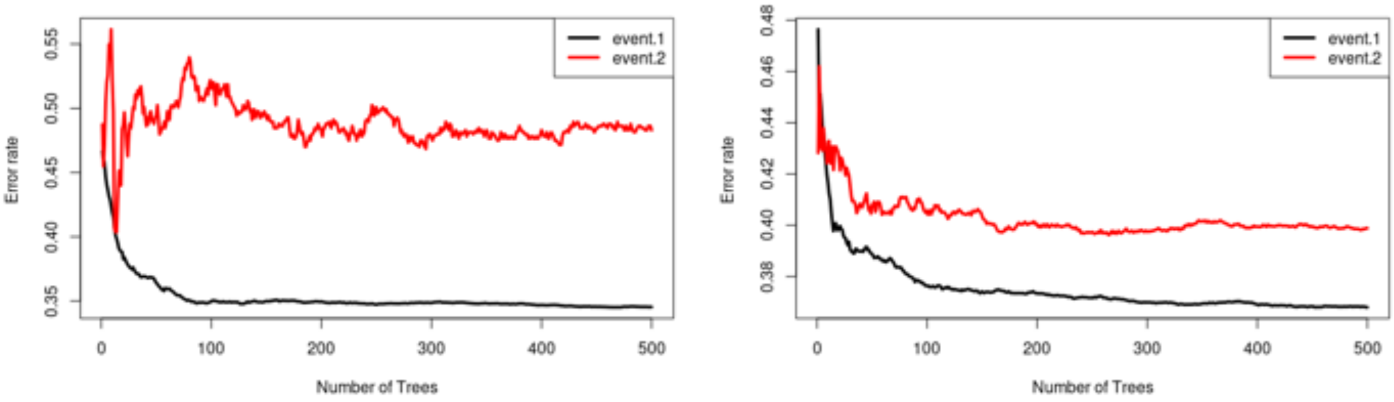

Figure S3. Partial variable dependence plots (with loess smoothing) showing the association of age (x-axis) with the cumulative incidence function at 60 days post-admission (y-axis) for discharge (left) and death (right) as computed from the competing risk Random Survival Forests model with all the variables on the entire population.

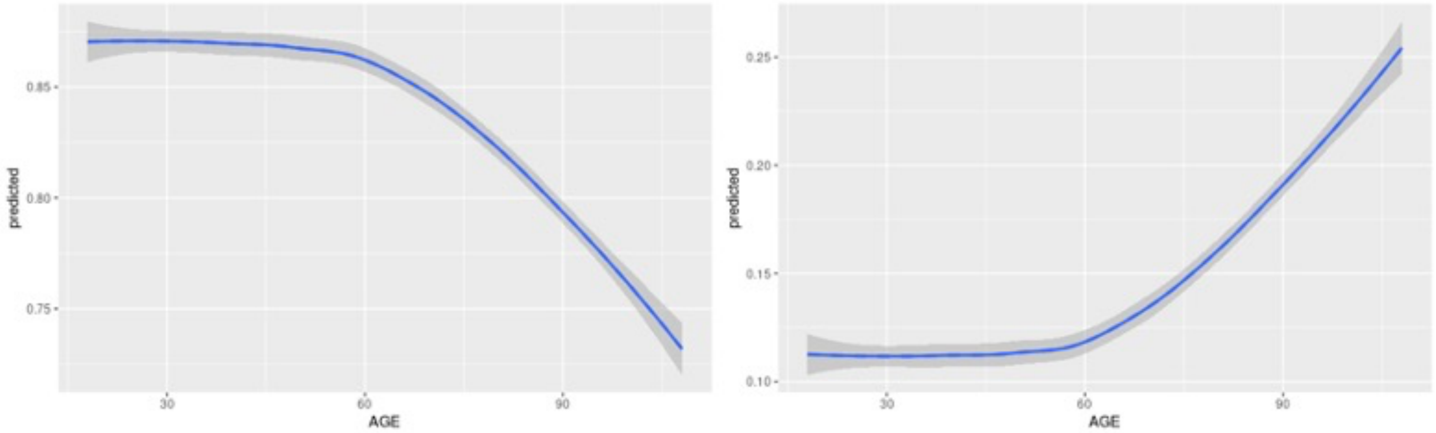

### *Variable importance calculated from RSF*

The most important predictors of hospital discharge and death are shown in Table S1 in descending order, starting from the top. Thirty among the 279 patients who died while hospitalized in one of the five JHHS hospitals were younger than 60 years. As a consequence, the predictive models for death among patients younger than 60 years should be interpreted with caution.

The relative importance for all variables across different domains pertaining to hospital discharge and mortality for patients 60 years and older (top) as well as to hospital discharge among those younger than 60 years of age (bottom). When compared to room air breathing, magnitude of oxygen requirement expressed as flow rate, device complexity and invasiveness required to deliver acceptable oxygenation progressively reduced the likelihood and time to hospital discharge among older and younger patients with COVID-19 and predicted death among those aged 60 and older. Lower respiratory and heart rates, lower BUN and procalcitonin levels, lesser inflammation as indexed by lower CRP levels as well as reduced red cell distribution width (RDW) were associated with greater likelihood and shortened time to hospital discharge in both the 60 and older and younger than 60 years of age with opposite directionality for incident death among those 60 and older. Other measures featured prominently on the Table S1 lists and Figure S4 plots for both age groups included eosinophil count, lymphocyte count, greater LDH and creatinine levels which were associated with a greater chance of death and lower chance of hospital discharge. Sepsis defined based on the systemic inflammatory response syndrome (SIRS) was clinically recognized by the presence of two or more of the following: (1)Temperature >38ºC or <36ºC, (2) Heart rate >90 beats/min, (3) Respiratory rate >20 breaths/min or PaCO2 <32 mmHg, (4) WBC >12,000 cells/mm3, <4000 cells/mm3, or >10 percent immature (band) forms. Sepsis was associated with a lower probability of discharge and a greater probability of death. Supplemental figures S3-S6 show partial effect plots from the RSF model used to understand non-linear associations and directionality.

Tissue damage indexed by higher levels of LDH was associated with later discharge in both age groups. Higher levels of muscle injury as indexed by creatine kinase and liver injury as assessed by AST/ALT ratio were markers of death in the young and old. Higher levels of ferritin were associated with early discharge in the younger age group. Specific markers of late discharge in the older age group were sepsis, neutrophilia, and monocytopenia. Specific markers of death in the older age group were higher mean corpuscular volume and hypotension. Among cardiac markers of stress or injury, higher levels of proBNP and several ECG parameters including PR interval, ventricular and atrial rates, and QRS axis were associated with worse outcomes.

Table S1: Top-20 cause-specific important variables (VIMP) for prediction of death and discharge from the competing risk RSF in patients less than 60 years of age, and 60 years and over.

| **Patients 60 years and over** | | |
|---|---|---|
| Rank | **VIMP – discharge** | **VIMP- death** |
| 1 | Oxygen support device | Oxygen support device |
| 2 | C-reactive protein | Respiratory rate |
| 3 | Respiratory rate | Blood Urea Nitrogen |
| 4 | Time since pandemic start | Time since pandemic start |
| 5 | Eosinophil % | Age |
| 6 | Ventricular rate | Eosinophil % |
| 7 | Blood Urea Nitrogen | Ventricular rate |
| 8 | Pulse rate | Red Cell Distribution Width |
| 9 | Age | Mean Corpuscular Volume |
| 10 | Eosinophil absolute count | Pulse rate |
| 11 | Lactate Dehydrogenase | Creatine Kinase |
| 12 | Procalcitonin | AST/ALT ratio |
| 13 | Red Cell Distribution Width | Procalcitonin |
| 14 | Pro B-type Natriuretic peptide | Atrial rate |
| 15 | Neutrophil % | Creatinine |
| 16 | Monocytes % | C-reactive protein |
| 17 | Sepsis | PR interval |
| 18 | Atrial rate | Systolic Blood pressure |
| 19 | Lymphocytes % | Color of Urine |
| 20 | QRS axis | Alcohol use |
| **Patients under 60 years of age** | | |
| Rank | **VIMP – discharge** | **VIMP- death** |
| 1 | Oxygen support device | Blood Urea Nitrogen |
| 2 | Age | Respiratory rate |
| 3 | Blood Urea Nitrogen | Pulse rate |
| 4 | Respiratory rate | Procalcitonin |
| 5 | Time since pandemic start | Red cell distribution width |
| 6 | C-reactive protein | Time since pandemic start |
| 7 | Procalcitonin | Creatinine |
| 8 | Pro B-Type Natriuretic Peptide | Mean corpuscular hemoglobin concentration |
| 9 | Lymphocytes % | Appearance of urine |
| 10 | Pulse rate | D-dimer |
| 11 | Creatinine | Pro B-type Natriuretic Peptide |
| 12 | Glucose | Alcohol abuse |
| 13 | Ferritin | AST/ALT ratio |
| 14 | Lactate Dehydrogenase | Height |
| 15 | Eosinophil % | Platelet count |
| 16 | Ventricular rate | Bilirubin |
| 17 | Red cell distribution width | Creatine kinase |
| 18 | Glomerular Filtration rate | Lymphocyte % |
| 19 | Atrial rate | Potassium |
| 20 | Blood loss anemia | Blood in urine |

Figure S4: Cause-specific variable importance (VIMP) for discharge (left column) and death (right), for variables from multiple domains as calculated from the competing risk RSF for patients >=60 years of age (top row) and patients <60 (bottom row). LDH – lactate dehydrogenase, BUN – blood urea nitrogen, PCT – procalcitonin, CRP – C-reactive protein, EOSINOPCT – eosiniphil %, EOSINOABS – eosinophil absolute count, MCV – mean corpuscular volume, RDW – red cell distribution width, PROBNP – NTproB-type natriuretic peptide, Resp_rate – respiratory rate, O2_device – oxygen device support type, DaysSinceStart – time since start of the pandemic.

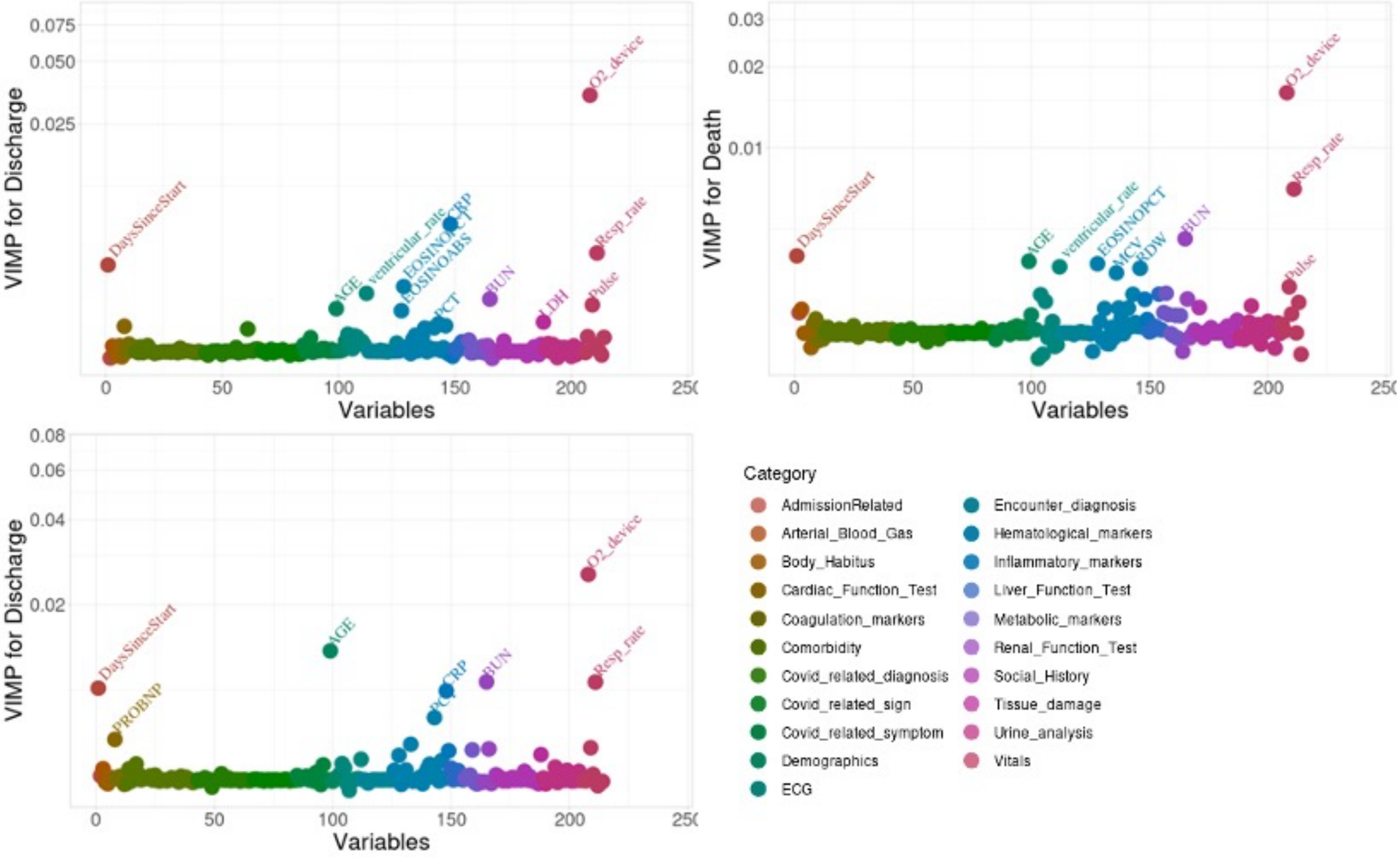

Figure S5: Partial plots for top-16 predictors of discharge from the competing risk RSF model in patients >=60 years. Y-axis denotes the CIF at 60 days. BUN (blood urea nitrogen), RDW (red cell distribution width), LDH (lactate dehydrogenase), PROBNP (proB-type natriuretic peptide), CRP (C-reactive protein) are log-transformed.

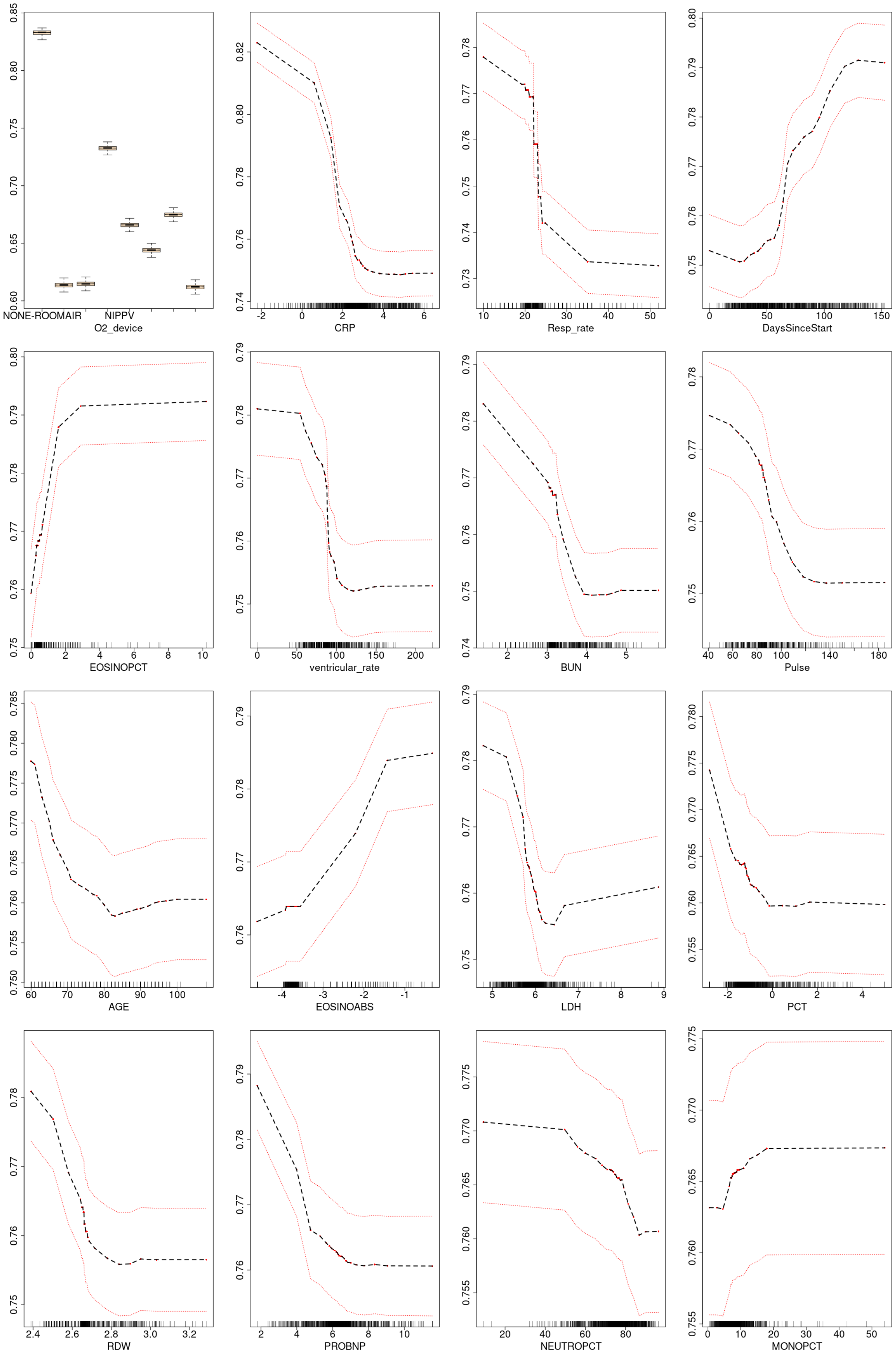

Figure S6: Partial plots for top-16 predictors of death from the competing risk RSF model in patients >=60 years. Y-axis denotes the CIF at 60 days. BUN (blood urea nitrogen), RDW (red cell distribution width), MCV (mean corpuscular volume), CKTOTAL (creatine kinase), ASTALT (AST/ALT ratio), PCT (procalcitonin), creatinine, CRP (C-reactive protein) are log-transformed.

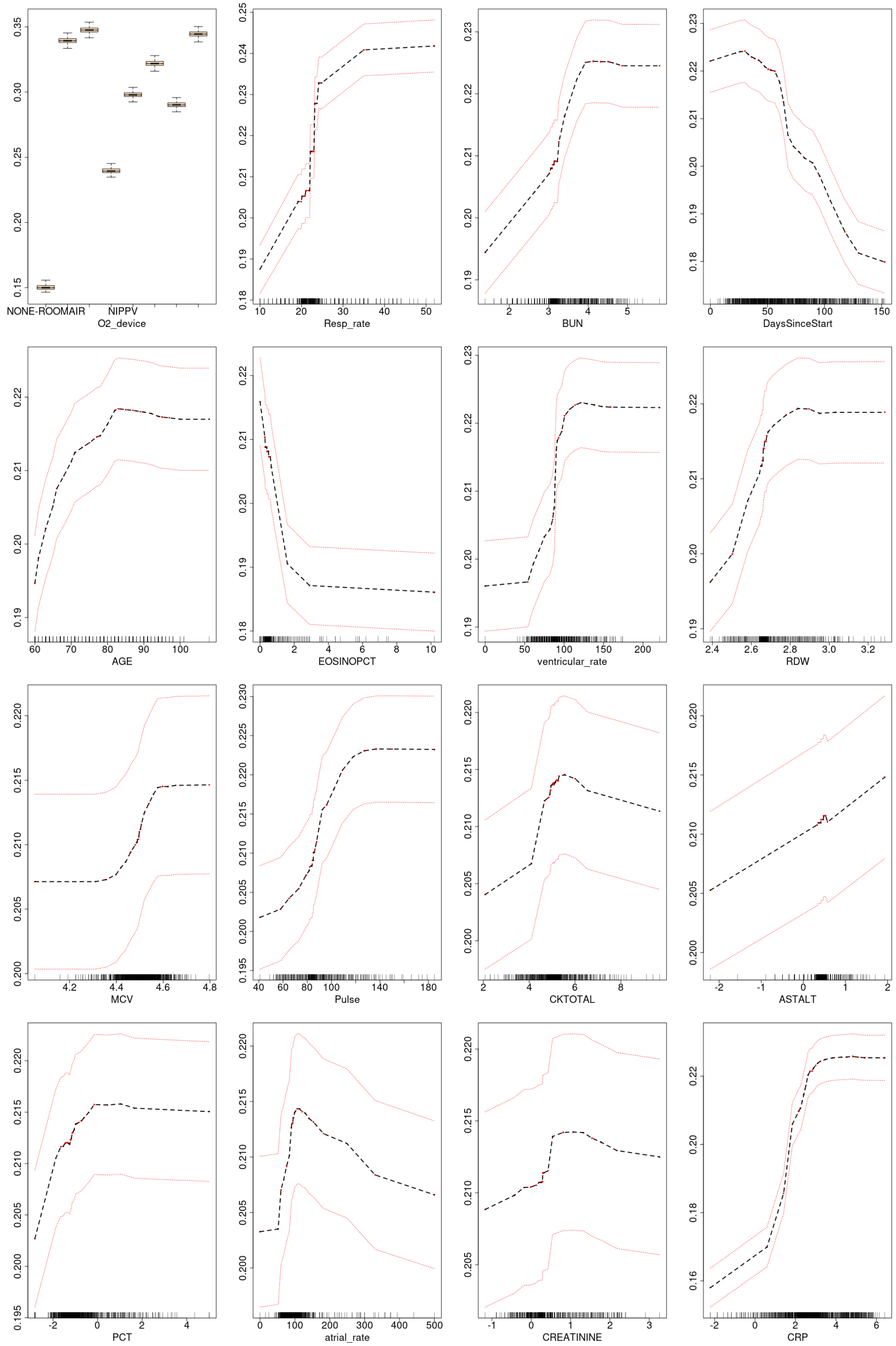

Figure S7: Partial plots for top-16 predictors of discharge from the competing risk RSF model in patients <60 years. Y-axis denotes the CIF at 60 days. BUN (blood urea nitrogen), LDH (lactate dehydrogenase), PROBNP (pro B-type natriuretic peptide), GLU (glucose), PCT (procalcitonin), creatinine, albumin, ferritin, and CRP (C-reactive protein) are log-transformed.

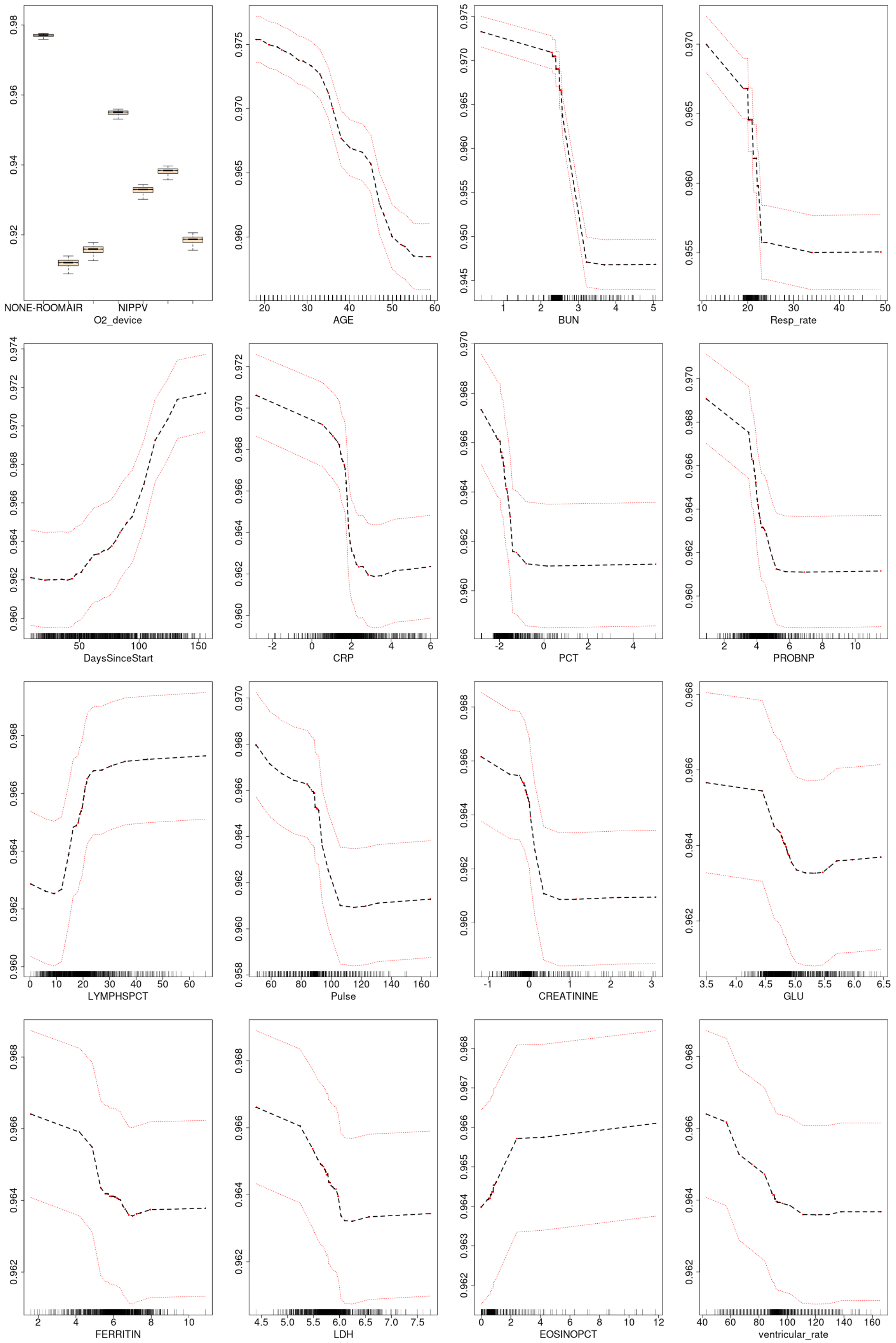

Figure S8: Partial plots for top-16 predictors of death from the competing risk RSF model in patients <60 years. Y-axis denotes the CIF at 60 days. BUN (blood urea nitrogen), PCT (procalcitonin), RDW (red cell distribution width), MCHC (mean hemoglobin corpuscular concentration), D-dimer, creatinine, AST/ALT ratio, PLT (platelet count), and BILITOT (bilirubin) are log-transformed

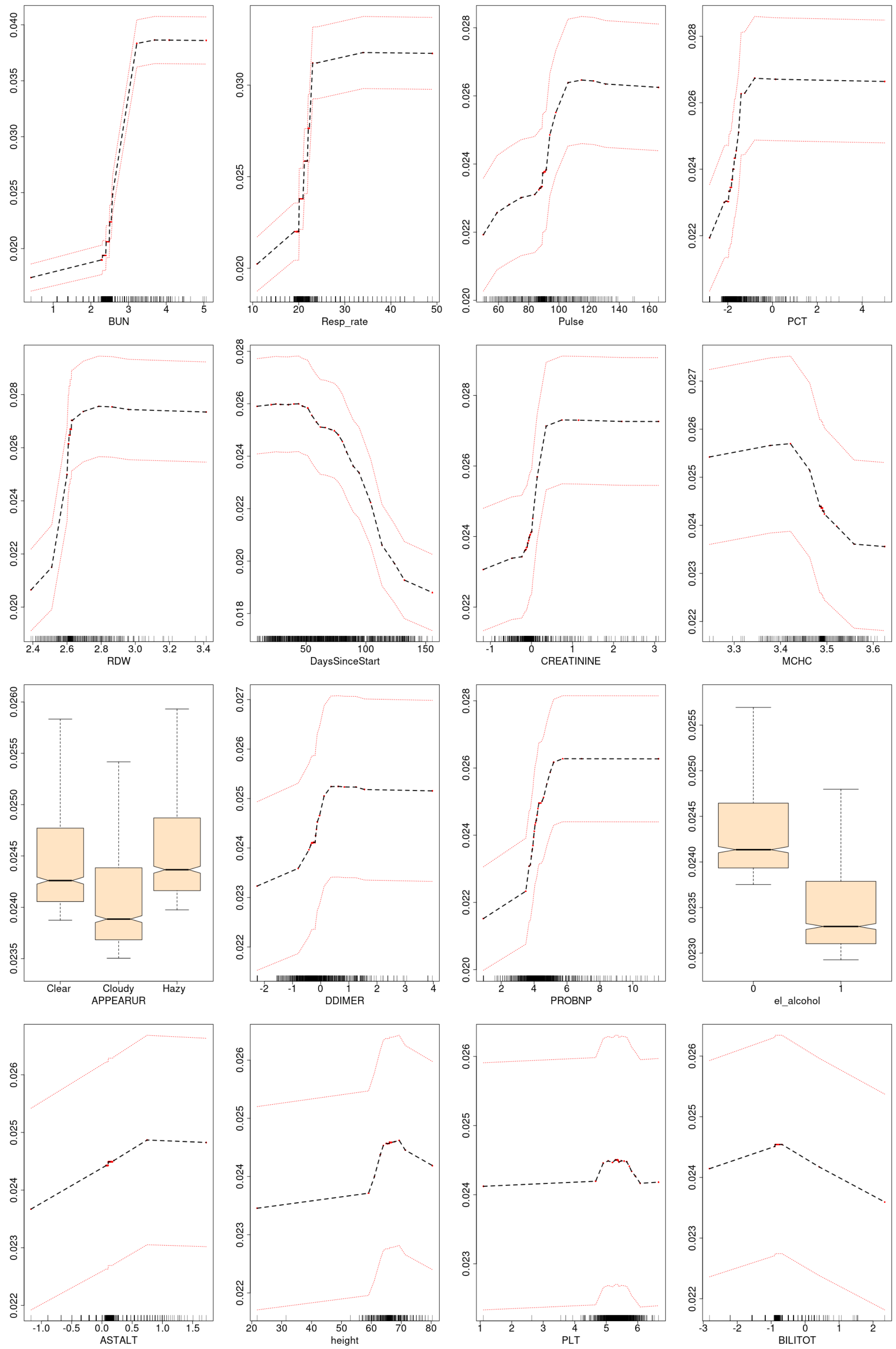

Figure S9: Model development was performed on a randomly chosen training subset (70% of the population) and applied on the test subset (remaining 30%) to ensure model calibration, validity and performance against the RSF models. Calibration plots for the Fine-Gray regression models showing the association (Loess curves) between predicted risk (x-axis) and observed event frequency (y-axis) at 20 days after admission using the test subset. The models were constructed on the training subset. Plots shown are for discharge (left column) and death (right), for patients >=60 years of age (top row) and patients <60 (bottom row). When oxygen device was present in the model, calibration was confirmed on models with and without this variable. The associated area under the curve (AUC) at 20 days post-admission is also shown. * - The number of deaths in the younger age group was n=30, appropriate care should be taken when interpreting the results.

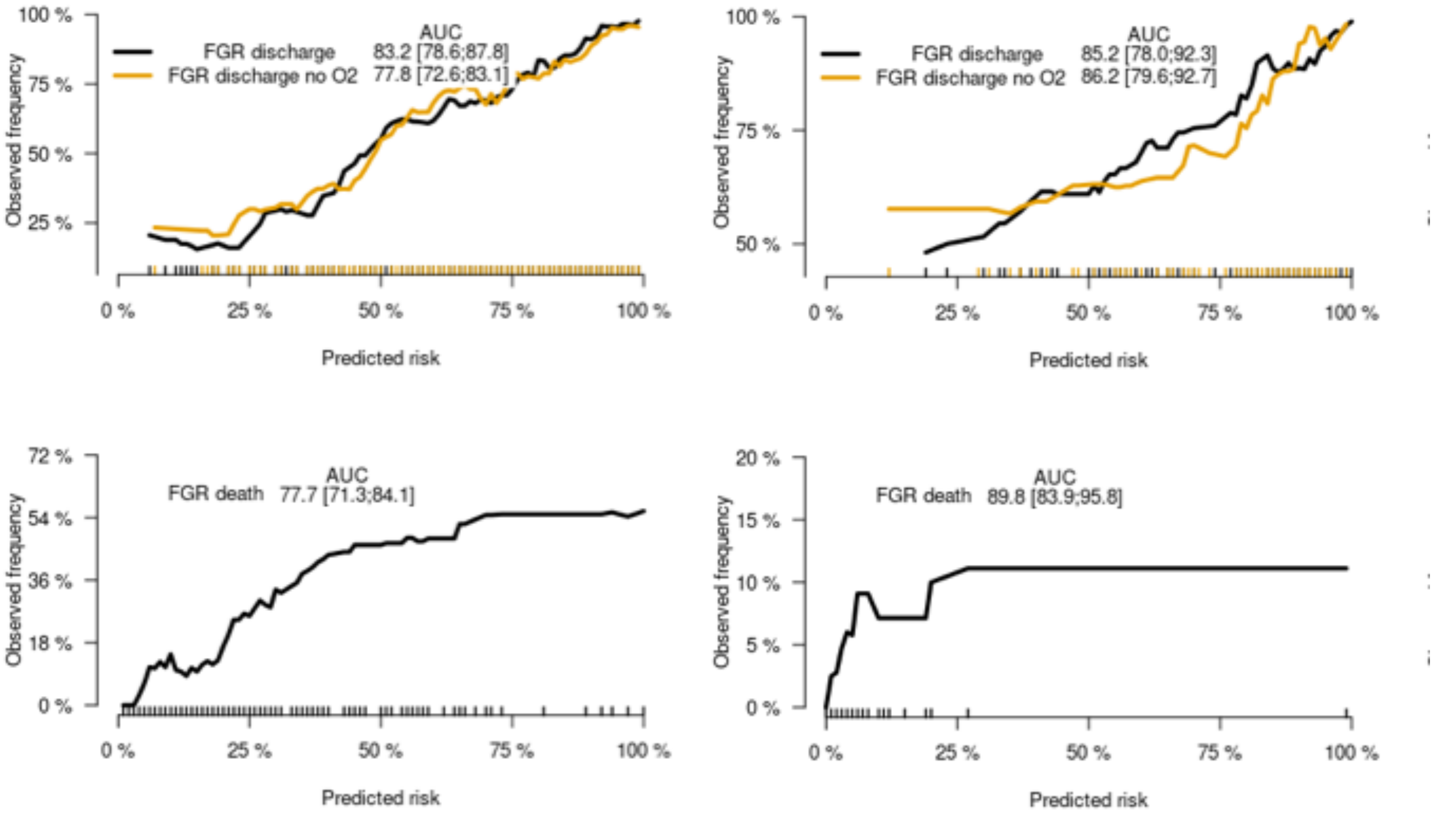

Figure S10: Model development was performed on a randomly chosen training subset (70% of the population) and applied on the test subset (remaining 30%) to ensure model calibration, validity and performance against the RSF models. Time-dependent area under the ROC for discharge (left panels) and death (right panels) from the competing risk Fine-Gray regression (FGR) and RSF models in patients 60 years and over (top row) and patients under 60 (bottom row) on the 'test' dataset. All models were constructed on 70% of the population (training data) and the results shown above generated for the same models on the remaining 30% of the population (testing data). The population was randomly divided into the testing and training data.
* - The number of deaths in the younger age group was n=30, appropriate care should be taken when interpreting the results.

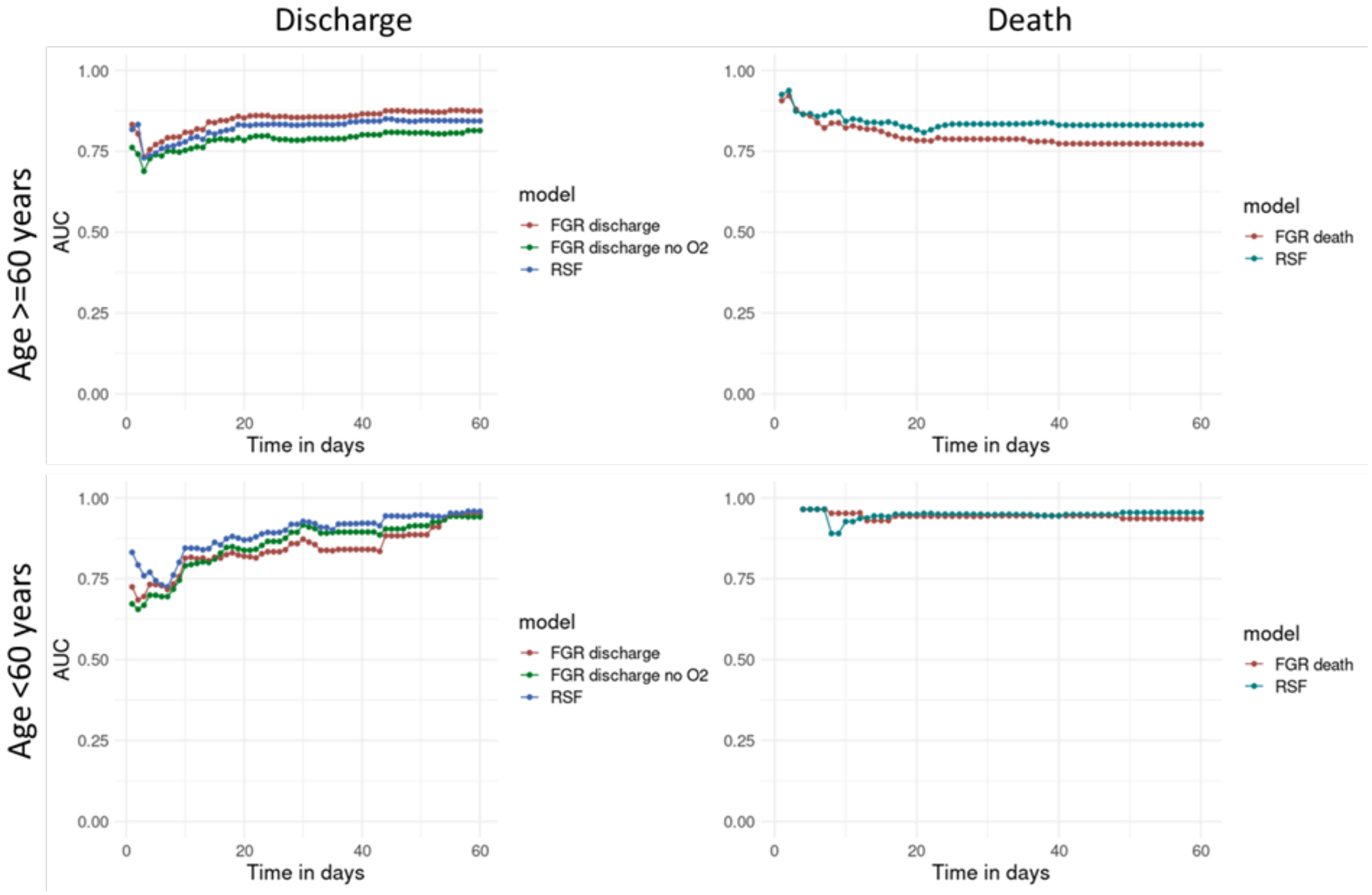
